# Body composition and checkpoint inhibitor treatment outcomes in advanced melanoma: a multicenter cohort study

**DOI:** 10.1101/2024.03.01.24303607

**Authors:** L.S. Ter Maat, I.A.J. Van Duin, R.J. Verheijden, P. Moeskops, J.J.C. Verhoeff, S.G. Elias, W.A.C. van Amsterdam, F.H. Burgers, F.W.P.J. Van den Berkmortel, M.J. Boers-Sonderen, M.F. Boomsma, J.W. De Groot, J.B.A.G. Haanen, G.A.P. Hospers, D. Piersma, G. Vreugdenhil, H.M. Westgeest, E. Kapiteijn, M. Labots, W.B. Veldhuis, P.J. Van Diest, P.A. De Jong, J.P.W. Pluim, T. Leiner, M. Veta, K.P.M. Suijkerbuijk

**Affiliations:** Image Sciences Institute, University Medical Center Utrecht, Utrecht University, Utrecht, The Netherlands; Department of Medical Oncology, University Medical Center Utrecht, Utrecht University, Utrecht, The Netherlands; Quantib, Rotterdam, The Netherlands; Department of Radiotherapy, University Medical Center Utrecht, Utrecht University, Utrecht, The Netherlands; Department of Epidemiology, Julius Center for Health Sciences and Primary Care, University Medical Center Utrecht, Utrecht University, Utrecht, The Netherlands; Department of Data Science and Biostatistics, Julius Center for Health Sciences and Primary Care, University Medical Center Utrecht, Utrecht University, Utrecht, The Netherlands; Department of Medical Oncology, Netherlands Cancer Institute, Amsterdam, the Netherlands; Department of Medical Oncology, Zuyderland Medical Center, Sittard-Geleen, The Netherlands; Department of Medical Oncology, Radboudumc, Radboud University, Nijmegen, The Netherlands; Department of Radiology, Isala Zwolle, Zwolle, The Netherlands; Isala Oncology Center, Isala Zwolle, Zwolle, The Netherlands; Department of Medical Oncology, Netherlands Cancer Institute, Amsterdam, The Netherlands; Department of Medical Oncology, Leiden University Medical Center, Leiden, The Netherlands; Melanoma Clinic, Centre Hospitalier Universitaire Vaudois, Lausanne, Switzerland; Department of Medical Oncology, UMC Groningen, University of Groningen, Groningen, The Netherlands; Department of Medical Oncology, Medisch Spectrum Twente, Enschede, The Netherlands; Department of Medical Oncology, Maxima Medical Center, Veldhoven, The Netherlands; Department of Internal Medicine, Amphia Hospital, Breda, The Netherlands; Department of Medical Oncology, Leiden University Medical Center, Leiden University, Leiden, The Netherlands; Department of Medical Oncology, Amsterdam UMC, Vrije Universiteit Amsterdam, Cancer Center Amsterdam, Boelelaan 1117, Amsterdam, The Netherlands; Department of Radiology, University Medical Center Utrecht, Utrecht University, Utrecht, The Netherlands; Department of Pathology, University Medical Center Utrecht, Utrecht University, Utrecht, The Netherlands; Medical Image Analysis, Department of Biomedical Engineering, Eindhoven University of Technology, Eindhoven, The Netherlands; Department of Radiology, Mayo Clinical, Rochester, Minnesota, USA

## Abstract

**Introduction:** The association of body composition with checkpoint inhibitor outcomes in melanoma is a matter of ongoing debate. In this study, we aim to add to previous evidence by investigating body mass index (BMI) alongside CT derived body composition metrics in the largest cohort to date.

**Method:** Patients treated with first-line anti-PD1 ± anti-CTLA4 for advanced melanoma were retrospectively identified from 11 melanoma reference centers in The Netherlands. Age, sex, Eastern Cooperative Oncology Group performance status, serum lactate dehydrogenase, presence of brain and liver metastases, number of affected organs and BMI at baseline were extracted from electronic patient files. From baseline CT scans, five body composition metrics were automatically extracted: skeletal muscle index, skeletal muscle density, skeletal muscle gauge, subcutaneous adipose tissue index and visceral adipose tissue index. All predictors were correlated in uni- and multivariable analysis to progression-free, overall and melanoma-specific survival (PFS, OS and MSS) using Cox proportional hazards models.

**Results:** A total of 1471 eligible patients were included. Median PFS and OS were 8.8 and 34.8 months, respectively. A significantly worse PFS was observed in underweight patients (multivariable HR=1.87, 95% CI 1.14–3.07). Furthermore, better OS was observed in patients with higher skeletal muscle density (multivariable HR=0.91, 95% CI 0.83-0.99) and gauge (multivariable HR=0.88, 95% CI 0.84-0.996), and a worse OS with higher visceral adipose tissue index (multivariable HR=1.13, 95% CI 1.04-1.22). No association with survival outcomes was found for overweightness or obesity and survival outcomes, or for subcutaneous adipose tissue.

**Discussion:** Our findings suggest that underweight BMI is associated with worse PFS, whereas higher skeletal muscle density and lower visceral adipose tissue index were associated with better OS. These associations were independent of previously identified predictors, including sex, age, performance status and extent of disease. No significant association between higher BMI and survival outcomes was observed.

## Introduction

The introduction of checkpoint inhibitors has revolutionized advanced melanoma care. The prognosis for advanced melanoma was historically very poor, with a 1-year overall survival of less than 25% [1]. In contrast, patients treated in the CheckMate 067 trial with anti-programmed cell death 1 (anti-PD1) had a 6.5-year overall survival rate of 43%. Patients treated with both anti-PD1 and anti-cytotoxic T-lymphocyte associated protein-4 (anti-CTLA4) antibodies even had a 6.5-year overall survival rate of 57% [2].

However, many open questions remain about how checkpoint inhibitors interact with tumor and host. Both anti-CTLA4 and anti-PD1 antibodies block proteins that inhibit immune response, which leads to increased immune activity against the tumor [3]. Although some mechanisms of primary resistance have been identified [4], it is not fully understood why some patients progress during treatment while others do not.

One such open question is the association between obesity and checkpoint inhibitor treatment outcomes. On the one hand, several pan-cancer meta-analyses published in 2020 and 2021 reported better survival outcomes in patients with obesity compared to patients with normal body mass index (BMI) [5–7]. This association, dubbed the “obesity paradox”, was also found to be significant in the subgroup of studies on patients with melanoma [6,7]. On the other hand, an updated meta-analysis by Roccuzzo et al. (2023) in melanoma concluded that the prognostic value of BMI could not be confirmed due to the limited available evidence [8]. This indicates that the topic of obesity and checkpoint inhibitor treatment outcomes is an area of ongoing research where more high-quality evidence is needed.

In addition to BMI, previous works investigated computed tomography (CT) derived body composition metrics. These metrics include the amount and density of skeletal muscle and the amount of subcutaneous and adipose tissue [9]. Due to advances in deep learning for automatic image analysis, this category of predictors has become increasingly prominent in research in recent years [10,11]. The advantage of these metrics is that they can more accurately capture a patient’s body composition, whereas BMI may misrepresent patients with high muscle mass and cannot distinguish between patients with high visceral or subcutaneous adipose tissue. Previous studies on these metrics, however, reported differing results and have some methodological limitations, most notably a limited sample size [12].

Several causal mechanisms have been proposed for explaining associations between body composition and checkpoint inhibitor outcomes. First, a more aggressive disease may affect both body composition (e.g., through weight loss) and outcomes. Second, patients with a worse physical condition, as reflected in body composition metrics, may succumb more quickly to their disease. Third, body composition may modulate the efficacy of checkpoint inhibitor therapy. For example, an increased efficacy of anti-PD(L)1 therapy was observed in obese mice compared to mice with normal weight [13]. Furthermore, increased PD-1 expression was noted in obese patients with melanoma [13].

Research into these causal mechanisms, however, is hindered by the controversy surrounding the association between body composition and checkpoint inhibitor treatment outcomes. This work therefore aimed to contribute to the existing evidence on this topic by presenting the largest cohort to our knowledge to date. Additionally, we aimed to provide a more fine-grained picture of body composition by evaluating CT derived metrics alongside BMI.

## Methods

### Patient selection

Patients were eligible if they were (i) over 18 years of age, (ii) treated for unresectable stage IIIC or stage IV cutaneous melanoma with (iii) first-line anti-PD1 with or without CTLA4 inhibition (iv) between January 1^st^, 2016, and February 1^st^, 2023. Patients were excluded if (i) no baseline CT scan was available, (ii) no transverse slice of the third lumbar vertebrae was in the field of view of the scan, (iii) metal artefacts were present at the L3 level or (iv) patient height or weight at baseline were unavailable. Eligible patients from eleven melanoma treatment centers in the Netherlands (Amphia Breda, Amsterdam UMC, Isala Zwolle, Leiden University Medical Center, Maxima MC, Medisch Spectrum Twente, Netherlands Cancer Institute, Radboudumc, University Medical Center Groningen, University Medical Center Utrecht, Zuyderland) were identified using high-quality registry data. This study was deemed not subject to Medical Research Involving Human Subjects Act according to Dutch regulations by the Medical Ethics Committee; informed consent was waived.

### BMI and clinical predictors

Height and weight at baseline were extracted from electronic patient files and were used to calculate BMI. In addition, several previously identified clinical predictors of checkpoint inhibitor treatment outcomes in advanced melanoma were extracted. These were (i) Eastern Cooperative Oncology Group (ECOG) performance status, (ii) level of lactate dehydrogenase (LDH), presence of (iii) brain and (iv) liver metastases and (v) number of affected organs [14–17] (categories are shown in Figure 1).

**Figure 1.**
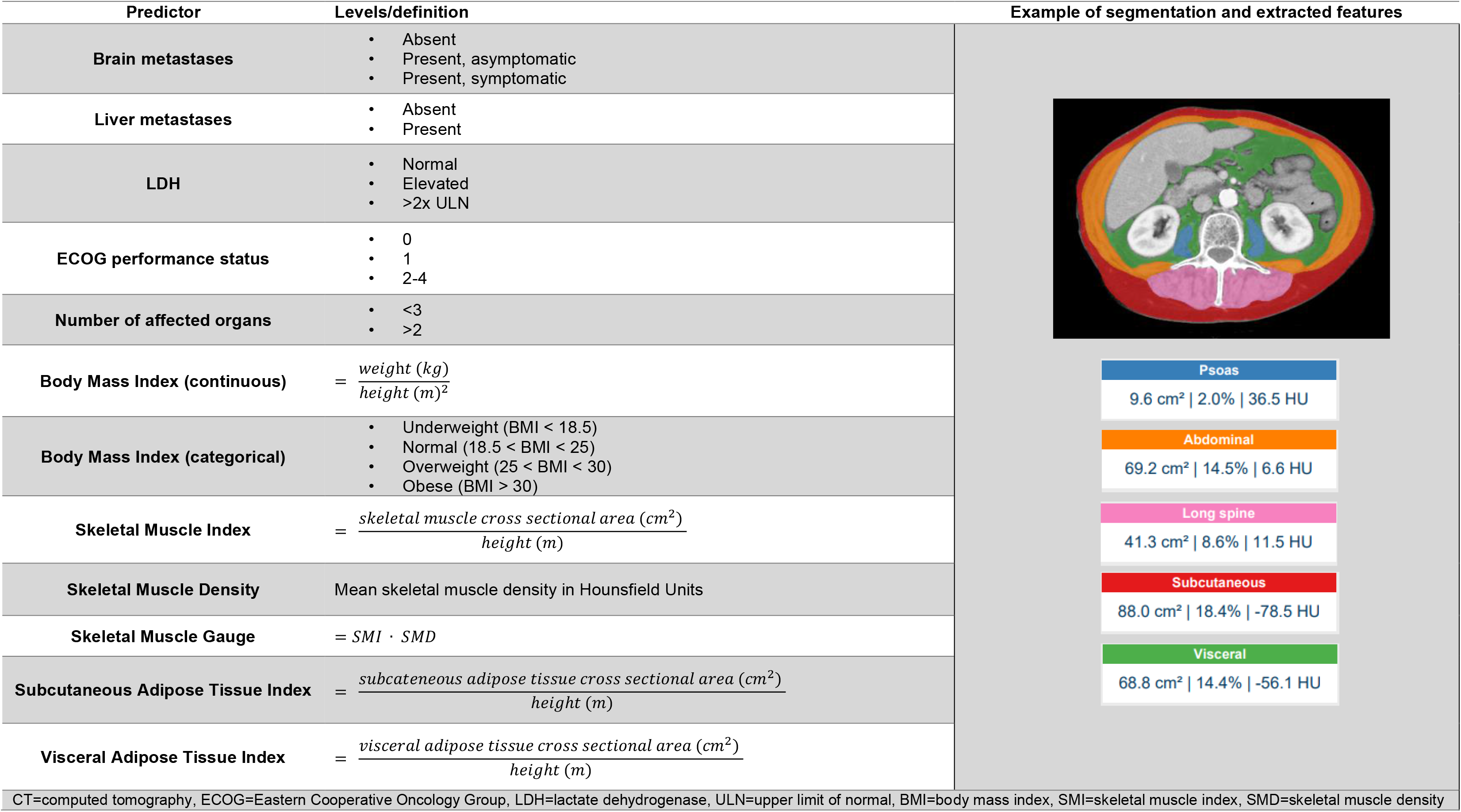
Definition of included predictors and evaluated models.

**Figure 2.**
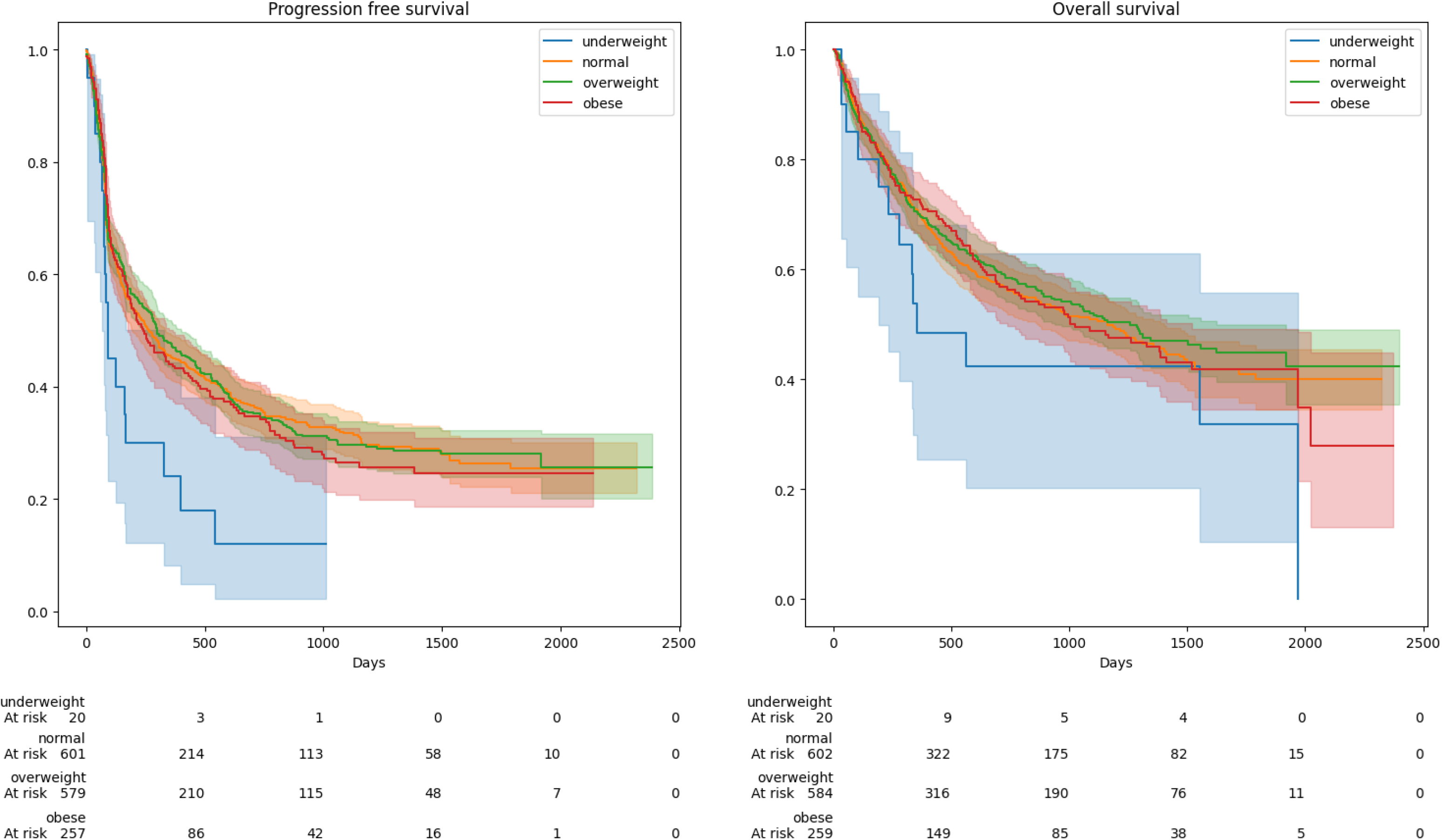
Kaplan-Meier curves for progression free and overall survival according to BMI subgroup.

### CT body composition metrics extraction

Metrics were obtained using Quantib Body Composition version 0.2.1, a dedicated deep learning segmentation algorithm that has proven to achieve high correspondence to manual segmentations in previous studies [18–20]. First, all baseline CT scans were resampled to a slice thickness of 5mm. Subsequently, the slice in the middle of the third lumbar vertebra [21] was automatically selected using a convolutional neural network. On the five consecutive slices centered around this selected slice, the following compartments were automatically segmented using a second convolutional neural network: psoas, abdominal and long spine muscles (together making up the skeletal muscles), subcutaneous adipose tissue and visceral adipose tissue. All segmentations were manually reviewed and corrected where necessary. Based on these segmentations, five commonly used metrics [9,22–24] were calculated using the definitions in Table 1: skeletal muscle index (SMI), skeletal muscle density (SMD), skeletal muscle gauge (SMG), subcutaneous adipose tissue index (SATI) and visceral adipose tissue index (VATI). All metrics were normalized to zero mean and unit standard deviation (SD) to facilitate interpretation. Since skeletal muscle density and gauge differed significantly between patients who underwent a contrast-enhanced CT scan versus those who underwent a non-contrast CT scan, SMD and SMG were normalized separately for both groups.

**Table 1.**
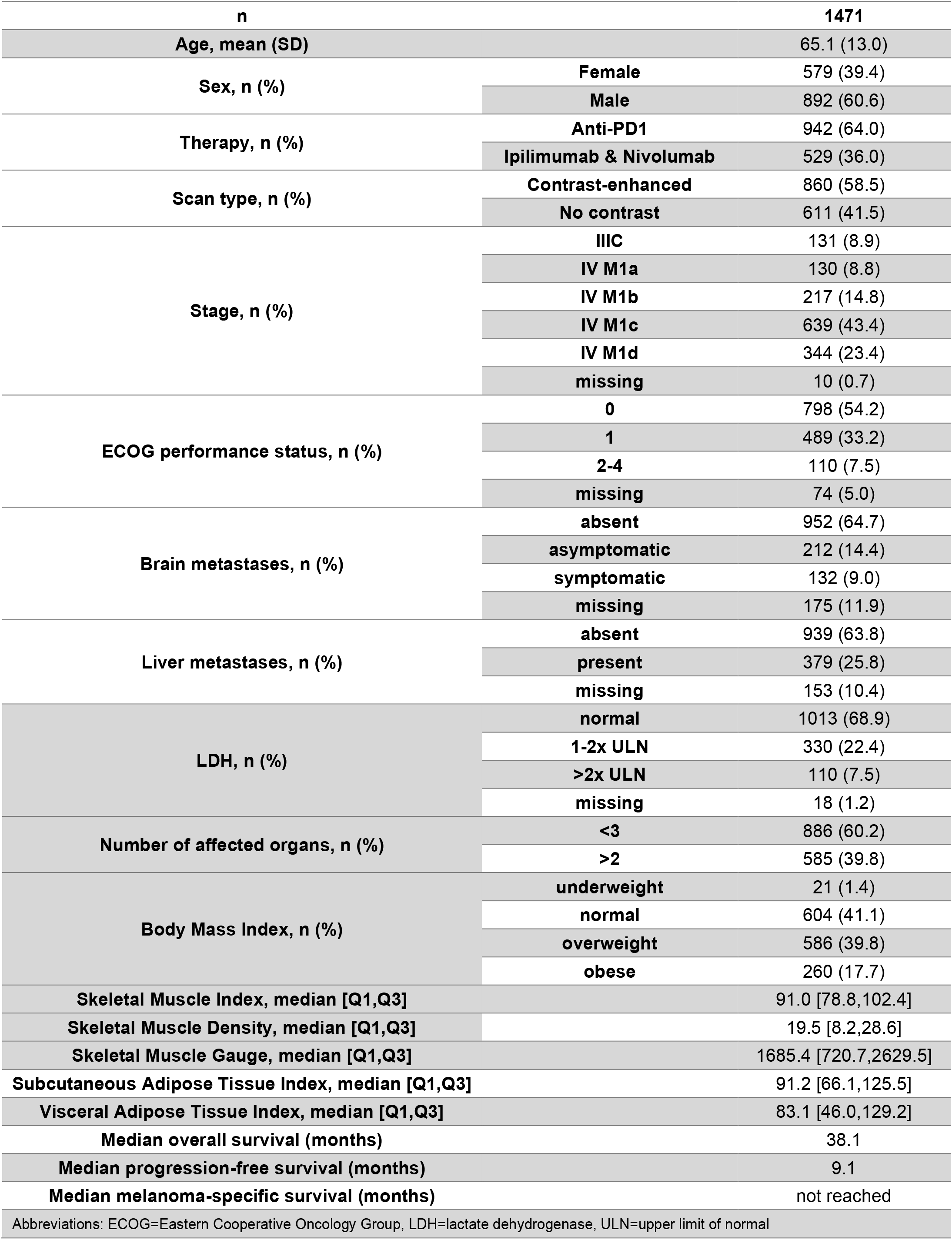
Characteristics of included patients.

| n |  | 1471 |
| --- | --- | --- |
| Age, mean (SD) |  | 65.1 (13.0) |
| Sex, n (%) | Female | 579 (39.4) |
|  | Male | 892 (60.6) |
| Therapy, n (%) | Anti-PD1 | 942 (64.0) |
|  | Ipilimumab & Nivolumab | 529 (36.0) |
| Scan type, n (%) | Contrast-enhanced | 860 (58.5) |
|  | No contrast | 611 (41.5) |
| Stage, n (%) | IIIC | 131 (8.9) |
|  | IV M1a | 130 (8.8) |
|  | IV M1b | 217 (14.8) |
|  | IV M1c | 639 (43.4) |
|  | IV M1d | 344 (23.4) |
|  | missing | 10 (0.7) |
| ECOG performance status, n (%) | 0 | 798 (54.2) |
|  | 1 | 489 (33.2) |
|  | 2-4 | 110 (7.5) |
|  | missing | 74 (5.0) |
| Brain metastases, n (%) | absent | 952 (64.7) |
|  | asymptomatic | 212 (14.4) |
|  | symptomatic | 132 (9.0) |
|  | missing | 175 (11.9) |
| Liver metastases, n (%) | absent | 939 (63.8) |
|  | present | 379 (25.8) |
|  | missing | 153 (10.4) |
| LDH, n (%) | normal | 1013 (68.9) |
|  | 1-2x ULN | 330 (22.4) |
|  | >2x ULN | 110 (7.5) |
|  | missing | 18 (1.2) |
| Number of affected organs, n (%) | <3 | 886 (60.2) |
|  | >2 | 585 (39.8) |
| Body Mass Index, n (%) | underweight | 21 (1.4) |
|  | normal | 604 (41.1) |
|  | overweight | 586 (39.8) |
|  | obese | 260 (17.7) |
| Skeletal Muscle Index, median [Q1,Q3] |  | 91.0 [78.8,102.4] |
| Skeletal Muscle Density, median [Q1,Q3] |  | 19.5 [8.2,28.6] |
| Skeletal Muscle Gauge, median [Q1,Q3] |  | 1685.4 [720.7,2629.5] |
| Subcutaneous Adipose Tissue Index, median [Q1,Q3] |  | 91.2 [66.1,125.5] |
| Visceral Adipose Tissue Index, median [Q1,Q3] |  | 83.1 [46.0,129.2] |
| Median overall survival (months) |  | 38.1 |
| Median progression-free survival (months) |  | 9.1 |
| Median melanoma-specific survival (months) |  | not reached |
| Abbreviations: ECOG=Eastern Cooperative Oncology Group, LDH=lactate dehydrogenase, ULN=upper limit of normal |  |  |

### Outcome definition

The primary endpoints were progression-free survival (PFS) and overall survival (OS). PFS was defined as the time from the start of treatment to progression or death; OS was defined as time from the start of treatment to death due to any cause. The secondary outcome was melanoma-specific survival (MSS), defined as the time from the start of treatment to death from melanoma. Patients not reaching the endpoint were right-censored at the date of the last contact, or when a different treatment was initiated.

### Statistical analysis

Correlation among body composition variables was assessed using Pearson’s correlation coefficient. The association between body composition metrics and outcomes were assessed using uni- and multivariable Cox proportional hazards models. In multivariable analyses, a separate model was constructed for every body composition metric, combined with previously identified clinical factors (ECOG performance status, level of LDH, presence of brain and liver metastases and number of affected organs). BMI was assessed as a categorical variable, using the established cut-offs for underweight (< 18.5), normal (between 18.5 and 25), overweight (between 25 and 30) and obese (>30). In addition, all variables were modelled using restricted cubic splines with three knots to account for non-linear effects. Multiple imputation was performed using the MICE R package with 21 imputations. Subgroup analyses were conducted for patients treated with monotherapy (anti-PD1) and combination therapy (anti-PD1 + anti-CTLA4), and for patients who underwent a contrast-enhanced and non-contrast CT scan. Unless stated otherwise, 95% confidence intervals are displayed.

## Results

### Patient characteristics

Out of 1944 eligible patients, 1471 patients (76%) were included (Supplementary Figure 1). Characteristics of the included patients are shown in Table 1; these characteristics were similar to those of excluded patients (Supplementary Table 1). Median PFS and OS were 9.1 and 38.1 months, respectively. Median MSS was not reached. The subgroups of patients treated with anti-PD1 monotherapy and anti-PD1 plus anti-CTLA-4 combination therapy consisted of 942 (64%) and 529 (36%) patients, respectively. Subgroups of patients who underwent non-contrast CT (in combination with 18-fluorodeoxyglucose positron emission tomography) versus contrast-enhanced consisted of 611 and 860 patients, respectively. Characteristics of patients in subgroups are shown in Supplementary Tables 2 and 3.

### Body mass index

Out of 1471 patients, 21 (1.4%) were underweight, 604 (41.1%) had normal BMI, 586 (39.8%) were overweight and 260 (17.7%) were obese. Underweight patients had significantly worse PFS than patients with normal weight in both uni- and multivariable analysis (multivariable HR=1.87 95% CI 1.14-3.07, Table 2, Figure 1). A similar, but statistically nonsignificant association was observed for OS (multivariable HR=1.57, 95% CI 0.89-2.77, Table 3, Figure 1). Underweight patients had more advanced disease, worse ECOG performance status, higher levels of LDH at baseline and were less likely to receive combination therapy (Supplementary Table 4). OS and PFS were not significantly different in overweight or obese patients when compared to normal BMI. No significant associations with OS and PFS were observed when BMI was analyzed using restricted cubic splines (Supplementary Figures 2-3). Results were comparable in the performed subgroup analyses (Supplementary Tables 6-13).

**Table 2.**
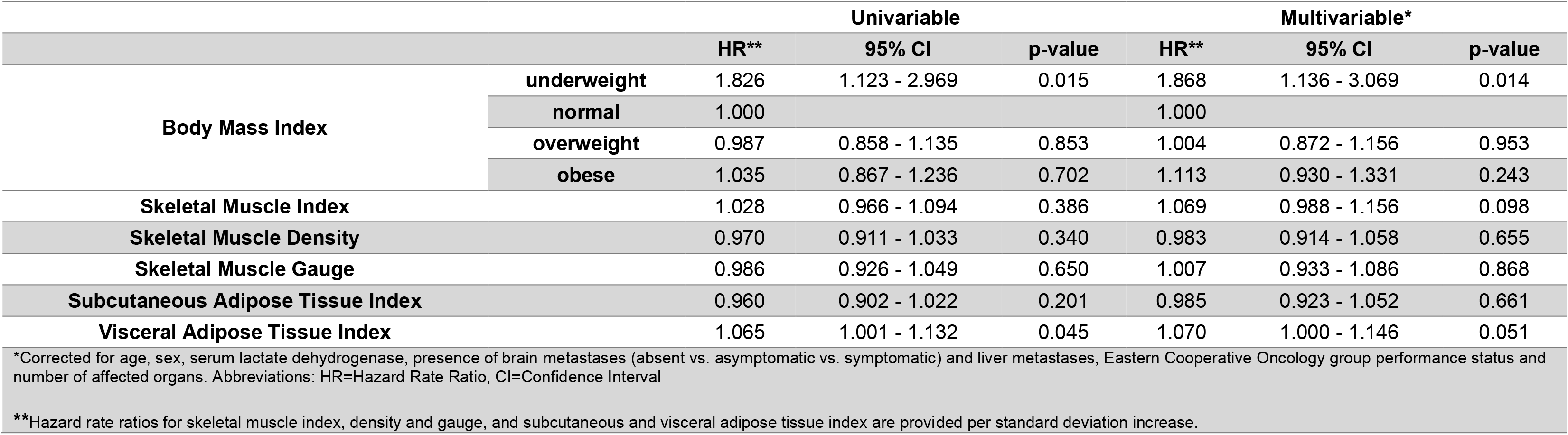
Univariable and multivariable Cox proportional hazards models for progression free survival.

**Table 3.**
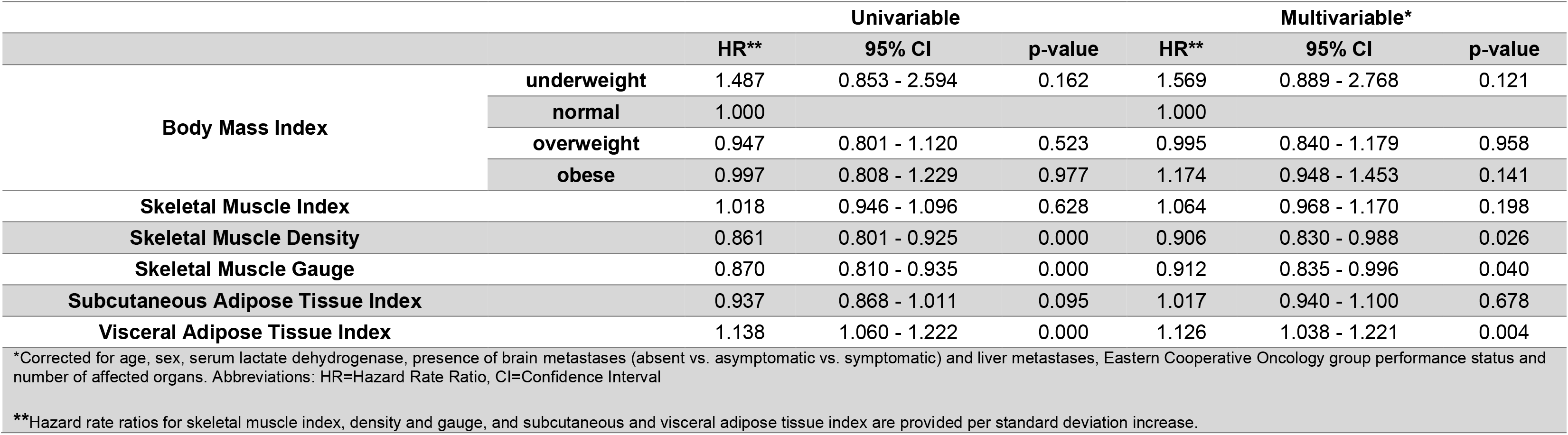
Univariable and multivariable Cox proportional hazards models for overall survival.

### CT derived body composition metrics

All body composition metrics were significantly correlated with each other (Supplementary Table 5). Of note is the negative correlation between skeletal muscle index and density (r = - 0.14). Significant associations with outcomes were observed for three of the five CT derived body composition metrics. First, higher skeletal muscle density was associated with better OS (multivariable HR=0.91 per SD increase, 95% CI 0.83-0.99, Table 3) and MSS (multivariable HR=0.90 per SD increase, 95% CI 0.81-0.999, Table 4). Second, higher skeletal muscle gauge was associated with better OS (multivariable HR=0.91 per SD increase, 95% CI 0.86-0.996, Table 3). Third, higher visceral adipose tissue index was associated with worse OS (multivariable HR=1.13 per SD increase, 95% CI 1.04-1.22, Table 3), with similar but statistically non-significant trends for PFS (multivariable HR=1.07 per SD increase, 95% CI 1.00-1.15, Table 2) and MSS (multivariable HR=1.10 per SD increase, 95% CI 0.997-1.21, Table 4). No significant associations were observed between skeletal muscle index or subcutaneous adipose tissue index and survival outcomes. Results were similar in subgroups of patients who underwent contrast-enhanced and non-contrast CT scans, in subgroups of patients treated with anti-PD1 and combination therapy (Supplementary Tables 6-13). When analyzing CT derived body composition metrics using restricted cubic splines, similar directions of effect were observed (Supplementary Figures 2-3).

**Table 4.**
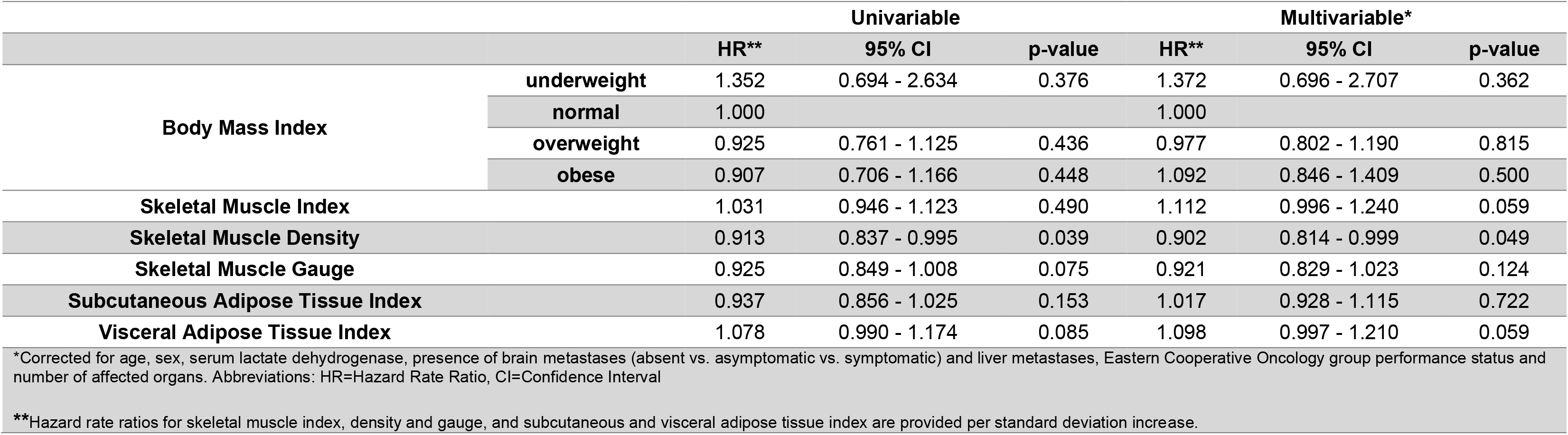
Univariable and multivariable Cox proportional hazards models for melanoma specific survival.

## Discussion

The contributions of this work are threefold. First, we demonstrate significantly worse PFS in patients who are underweight. Second, we find no evidence for an association between obesity and better outcomes. Third, we show that higher skeletal muscle density and gauge, and lower visceral adipose tissue index are associated with improved survival.

PFS was significantly worse in underweight patients. Surprisingly, this association was significant in multivariable analysis despite the association between underweight BMI and other poor baseline characteristics. Although this result must be interpreted with care due to the small numbers (N=21) in the underweight group, it may indicate that the prognosis of this group of patients is even worse than is to be expected based on their stage of disease, performance status and level of LDH. Potential explanations for this association are a confounding effect of tumor aggressiveness, and an increased vulnerability to complications due to reduced physical reserves.

We found no association between obesity and better treatment outcomes, when measured as BMI, or as visceral or subcutaneous adipose tissue index. In contrast, we observed worse survival in patients with more visceral adipose tissue, the type of fat most associated with inflammation [25]. The other metrics that reflect obesity, namely subcutaneous adipose tissue index and higher BMI, were not associated with any of the investigated outcomes. These findings are in line with the meta-analysis by Roccuzzo et al. [8], which found no significant association between higher BMI and survival outcomes in melanoma. This meta-analysis thereby differs in its conclusion from earlier meta-analyses, a fact which can be explained by the inclusion of studies which were not yet published during these earlier analyses.

Better survival was observed in patients with higher skeletal muscle density and gauge, and lower adipose tissue index. There are multiple explanations for the results. On the one hand, it could be that these metrics are general prognostic indicators irrespective of treatment. This interpretation is supported by the fact that the associations were stronger for overall survival than for PFS and MSS. On the other hand, it could be that body composition influences the effect of checkpoint inhibitor treatment. A proposed mechanism is that visceral adipose tissue dysregulates the body’s immune response, leading to worse treatment effects [26,27]. Future research, however, is needed to confirm this association and to determine the underlying causal mechanisms.

This study contributes to previous evidence in two important ways. First, it adds the largest cohort collected on this topic to date and thereby strengthens the conclusion of the meta-analysis by Roccuzzo et al. [8] regarding obesity. Second, it provides a more fine-grained view of body composition through the use of CT derived body composition metrics. This is particularly relevant in the case of visceral adipose tissue, where our findings suggest a negative association with survival, rather than a positive one as was suggested by earlier findings on BMI.

A limitation is the exclusion of otherwise eligible patients due to unavailable data. Approximately 25% of eligible patients were excluded due to lack of required data. We argue, however, that the risk of selection bias is limited, as differences in patient characteristics between included and excluded patients were small. Furthermore, the correction of skeletal muscle density for the presence of contrast is likely to be imperfect. This correction assumes that the mean and standard deviation of the true skeletal muscle density is the same for patients who underwent contrast-enhanced and no-contrast baseline scans. This may not be the case, given the difference in patient characteristics between the two groups. Given the consistent results in the subgroup analyses, we think it is unlikely that this imperfect correction would have significantly influenced the results.

In conclusion, underweight BMI, more visceral adipose tissue and lower skeletal muscle density are associated with worse outcomes in ICI treated advanced melanoma patients, independent of known predictors. The significance of the associations in multivariable analysis indicates that the information provided by body composition metrics is not fully captured by previously identified predictors, such as ECOG performance status. Outcomes were not significantly different in overweight and obese patients, as compared with those with normal BMI. This finding is in accordance with a recent meta-analysis on this topic. Our work contributes to previous research by presenting the largest cohort to date and by providing detailed data on body composition through CT derived metrics.

## Data Availability

Data produced in the present work is not available due to confidentiality agreements.

## Conflict of interest statement

AvdE has advisory relationships with Amgen, Bristol Myers Squibb, Roche, Novartis, MSD, Pierre Fabre, Sanofi, Pfizer, Ipsen, Merck and has received research study grants not related to this paper from Sanofi, Roche, Bristol Myers Squibb, Idera and TEVA and has received travel expenses from MSD Oncology, Roche, Pfizer and Sanofi and has received speaker honoraria from BMS and Novartis.

JdG has consultancy/advisory relationships with Bristol Myers Squibb, Pierre Fabre, Servier, MSD, Novartis.

GH has consultancy/advisory relationships with Amgen, Bristol-Myers Squibb, Roche, MSD, Novartis, Sanofi, Pierre Fabre and has received research grants from Bristol-Myers Squibb, Seerave. With all payments to the Institution.

PJ has a research collaboration with Philips Healthcare.

MBS has consultancy/advisory relationships with Pierre Fabre, MSD and Novartis.

EK has consultancy/advisory relationships with Bristol Myers Squibb, Delcath and Lilly,, and received research grants not related to this paper from Bristol Myers Squibb, Delcath, Novartis and Pierre Fabre. All paid to the institution.

PD has consultancy/advisory relationships with Paige, Visiopharm, Sectra, Pantarei and Samantree paid to the institution and research grants from Pfizer, none related to current work and paid to institute.

KS has advisory relationships with Pierre Fabre, AbbVie and Sairopa and received research funding from Bristol Myers Squibb, TigaTx, Philips and Genmab.

TL has received research funding from Philips. PM is employed at Quantib.

DP has advisory relationships with Novartis, Pierre Fabre en BMS and honorarium for lecturing from Novartis. Not related to current work.

All remaining authors have declared no conflicts of interest.

