## Supplementary Materials for "Body composition and checkpoint inhibitor treatment outcomes in advanced melanoma: a multicenter cohort study"

**Supplementary Figure 1 – Flowchart of the inclusion process**

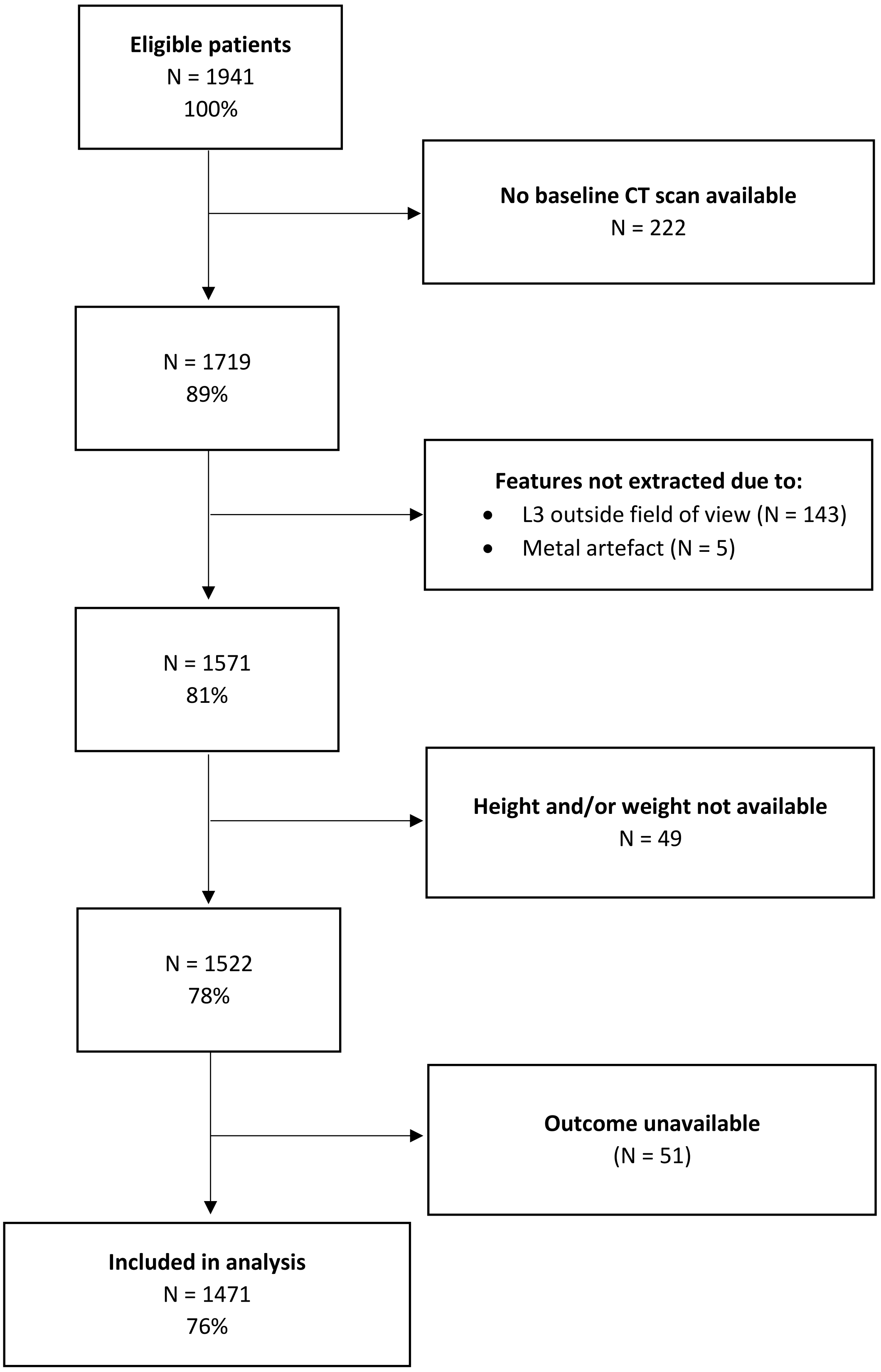

**Supplementary Figure 2 – Cox proportional hazards model regression with restricted cubic splines for progression free survival in univariable (left) and multivariable (right) analysis**

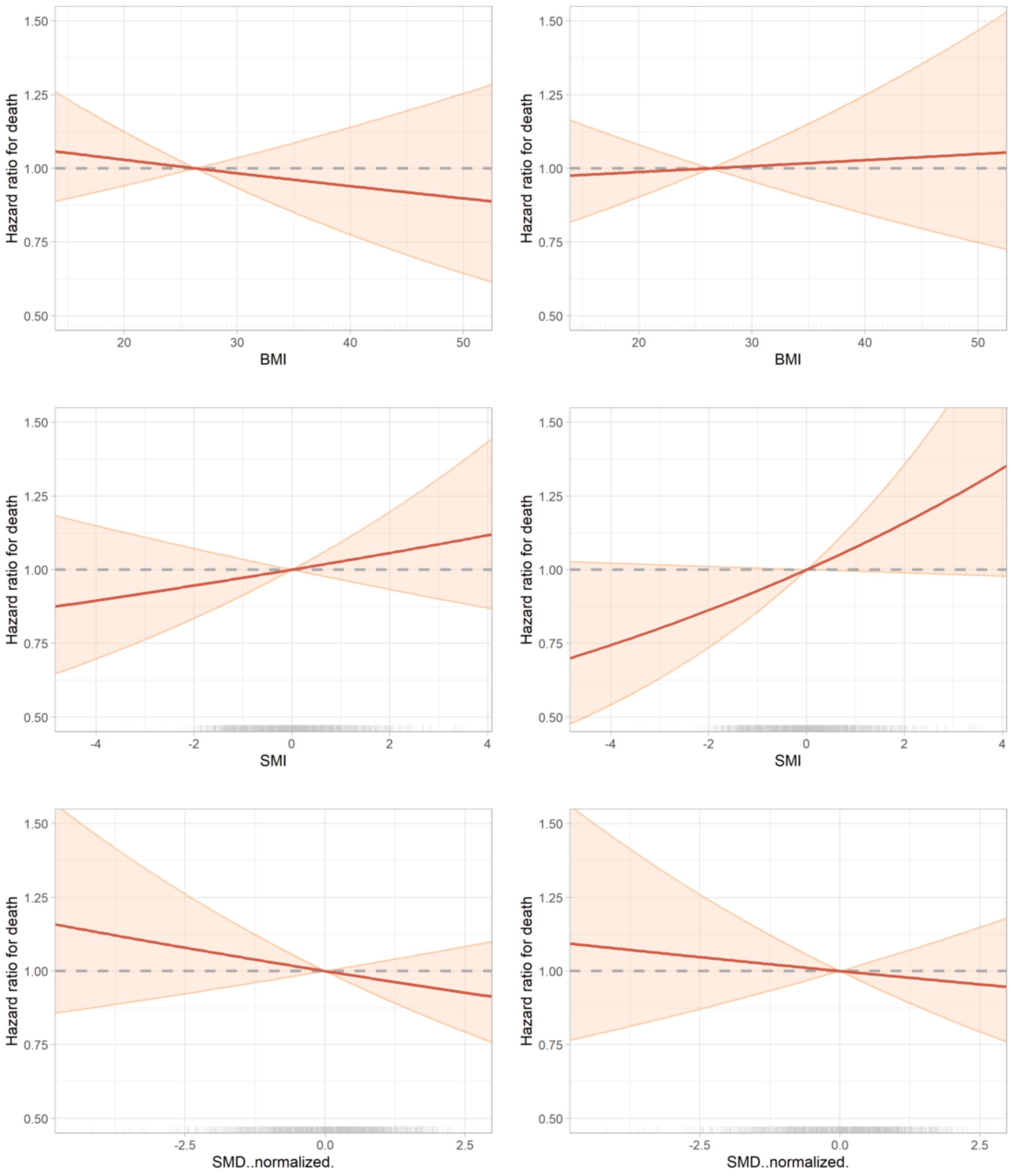

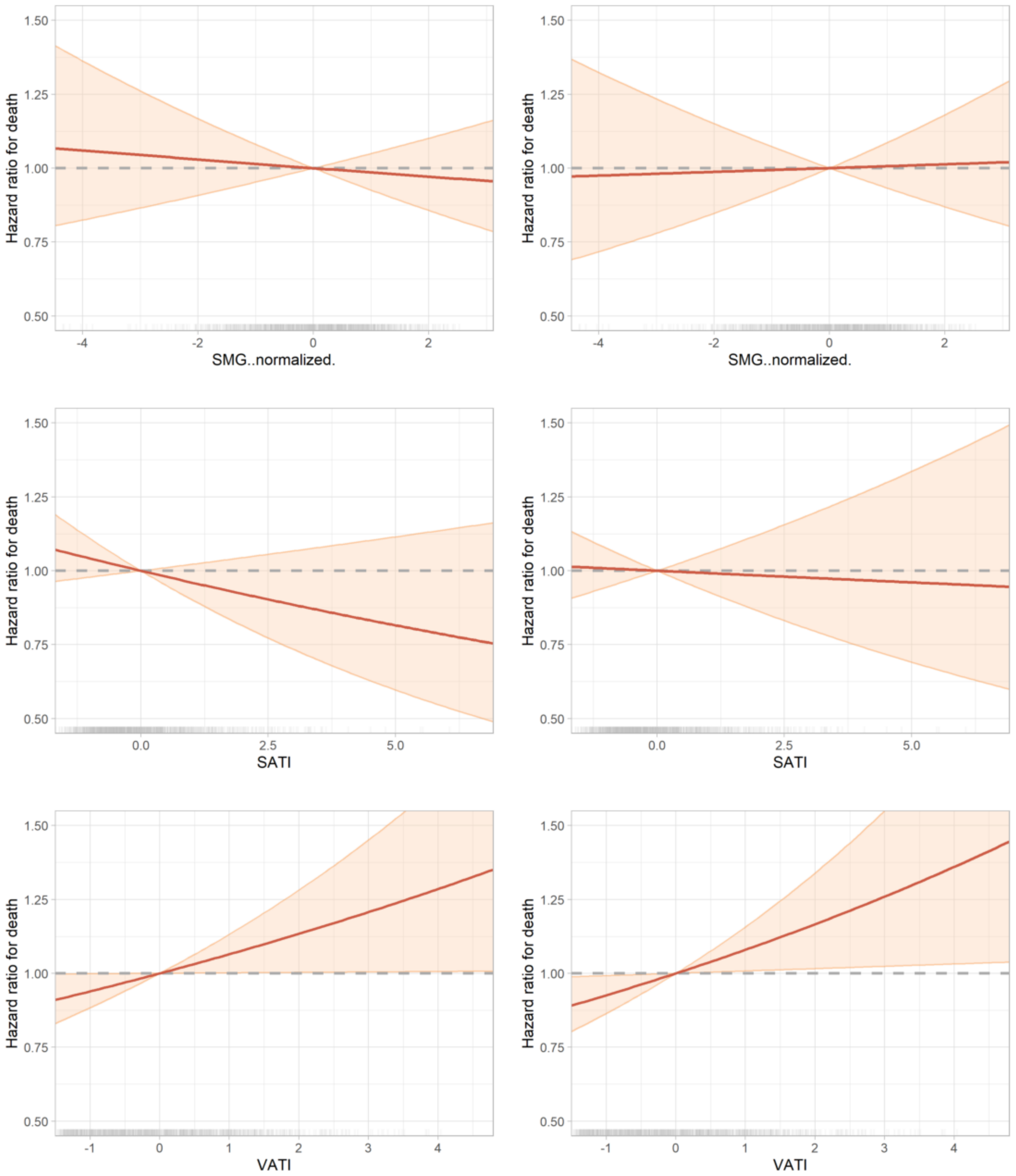
Hazard ratio for death
1.50
1.25
1.00
0.75
0.50
-4
-2
0
2
SMG..normalized.
0.0
2.5
5.0
SATI
-1
0
1
2
3
4
VATI

**Supplementary Figure 3 – Cox proportional hazards model regression with restricted cubic splines for overall survival in univariable (left) and multivariable (right) analysis**

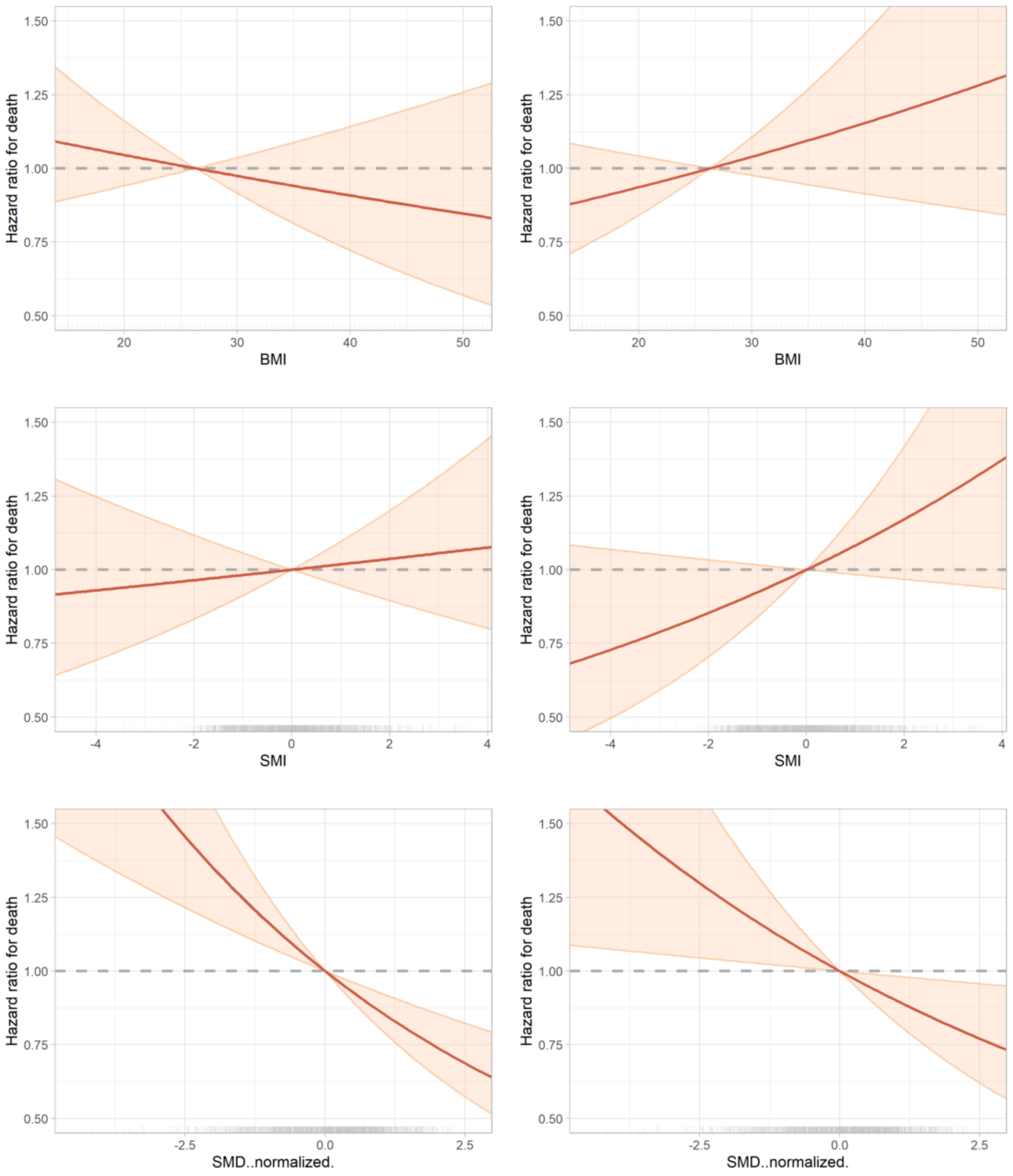

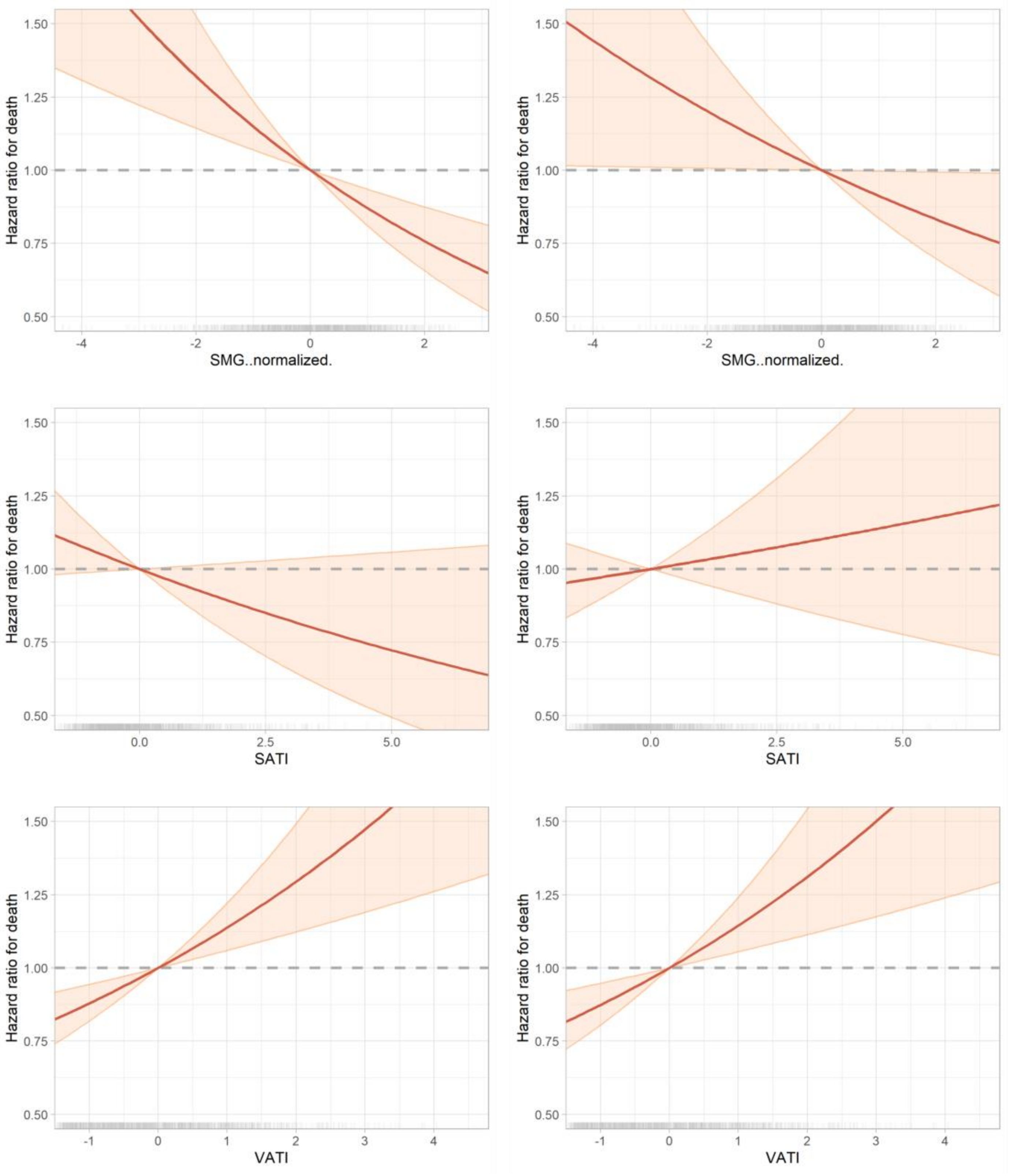
Hazard ratio for death
1.50
1.25
1.00
0.75
0.50
-4
-2
0
2
SMG..normalized.
Hazard ratio for death
1.50
1.25
1.00
0.75
0.50
-4
-2
0
2
SMG..normalized.
Hazard ratio for death
1.50
1.25
1.00
0.75
0.50
0.0
2.5
5.0
SATI
Hazard ratio for death
1.50
1.25
1.00
0.75
0.50
0.0
2.5
5.0
SATI
Hazard ratio for death
1.50
1.25
1.00
0.75
0.50
-1
0
1
2
3
4
VATI
Hazard ratio for death
1.50
1.25
1.00
0.75
0.50
-1
0
1
2
3
4
VATI

**Supplementary Table 1 - Characteristics and outcomes of included versus excluded patients**

| | | Included | Excluded |
|---|---|---|---|
| **n** | | 1471 | 470 |
| **Age, mean (SD)** | | 65.1 (13.0) | 65.0 (13.9) |
| **Sex, n (%)** | **Female** | 579 (39.4) | 167 (35.5) |
| | **Male** | 892 (60.6) | 303 (64.5) |
| **Therapy, n (%)** | **Anti-PD1** | 942 (64.0) | 300 (63.8) |
| | **Ipilimumab & Nivolumab** | 529 (36.0) | 170 (36.2) |
| **Scan type, n (%)** | **Contrast-enhanced** | 860 (58.5) | 53 (66.2) |
| | **No contrast** | 611 (41.5) | 27 (33.8) |
| **Stage, n (%)** | **IIIC** | 131 (8.9) | 33 (7.0) |
| | **IV M1a** | 130 (8.8) | 37 (7.9) |
| | **IV M1b** | 217 (14.8) | 77 (16.4) |
| | **IV M1c** | 639 (43.4) | 197 (41.9) |
| | **IV M1d** | 344 (23.4) | 119 (25.3) |
| | **missing** | 10 (0.7) | 7 (1.5) |
| **ECOG performance status, n (%)** | **0** | 798 (54.2) | 239 (50.9) |
| | **1** | 489 (33.2) | 177 (37.7) |
| | **2-4** | 110 (7.5) | 31 (6.6) |
| | **missing** | 74 (5.0) | 23 (4.9) |
| **Brain metastases, n (%)** | **absent** | 952 (64.7) | 308 (65.5) |
| | **asymptomatic** | 212 (14.4) | 66 (14.0) |
| | **symptomatic** | 132 (9.0) | 53 (11.3) |
| | **missing** | 175 (11.9) | 43 (9.1) |
| **Liver metastases, n (%)** | **absent** | 939 (63.8) | 314 (66.8) |
| | **present** | 379 (25.8) | 112 (23.8) |
| | **missing** | 153 (10.4) | 44 (9.4) |
| **LDH, n (%)** | **normal** | 1013 (68.9) | 302 (64.3) |
| | **1-2x ULN** | 330 (22.4) | 130 (27.7) |
| | **>2x ULN** | 110 (7.5) | 31 (6.6) |
| | **missing** | 18 (1.2) | 7 (1.5) |
| **Number of affected organs, n (%)** | **<3** | 886 (60.2) | 277 (58.9) |
| | **>2** | 585 (39.8) | 193 (41.1) |
| **Body Mass Index, n (%)** | **underweight** | 21 (1.4) | |
| | **normal** | 604 (41.1) | 30 (39.5) |
| | **overweight** | 586 (39.8) | 26 (34.2) |
| | **obese** | 260 (17.7) | 20 (26.3) |
| **Skeletal Muscle Index, median [Q1,Q3]** | | -0.0 [-0.7,0.6] | 0.1 [-0.5,0.6] |
| | | 0.1 [-0.6,0.7] | 0.1 [-0.7,0.7] |
| | | 0.1 [-0.6,0.7] | 0.1 [-0.8,0.5] |
| **Subcutaneous Adipose Tissue Index, median [Q1,Q3]** | | -0.2 [-0.7,0.4] | -0.5 [-0.9,0.2] |
| **Visceral Adipose Tissue Index, median [Q1,Q3]** | | -0.2 [-0.8,0.6] | -0.2 [-0.9,0.7] |
| **Median overall survival (months)** | | 38.1 | 29.1 |
| **Median progression-free survival (months)** | | 9.1 | 8.1 |
| **Median melanoma-specific survival (months)** | | not reached | 47.1 |

**Supplementary Table 2 - Characteristics and outcomes of patients who received anti-PD1 versus combination therapy**

| | | Anti-PD1 | Ipilimumab & Nivolumab |
|---|---|---|---|
| **n** | | 942 | 529 |
| **Age, mean (SD)** | | 67.0 (12.8) | 61.6 (12.6) |
| **Sex, n (%)** | **Female** | 380 (40.3) | 199 (37.6) |
| | **Male** | 562 (59.7) | 330 (62.4) |
| **Scan type, n (%)** | **Contrast-enhanced** | 538 (57.1) | 322 (60.9) |
| | **No contrast** | 404 (42.9) | 207 (39.1) |
| **Stage, n (%)** | **IIIC** | 113 (12.0) | 18 (3.4) |
| | **IV M1a** | 112 (11.9) | 18 (3.4) |
| | **IV M1b** | 184 (19.5) | 33 (6.2) |
| | **IV M1c** | 397 (42.1) | 242 (45.7) |
| | **IV M1d** | 130 (13.8) | 214 (40.5) |
| | **missing** | 6 (0.6) | 4 (0.8) |
| **ECOG performance status, n (%)** | **0** | 537 (57.0) | 261 (49.3) |
| | **1** | 291 (30.9) | 198 (37.4) |
| | **2-4** | 62 (6.6) | 48 (9.1) |
| | **missing** | 52 (5.5) | 22 (4.2) |
| **Brain metastases, n (%)** | **absent** | 672 (71.3) | 280 (52.9) |
| | **asymptomatic** | 80 (8.5) | 132 (25.0) |
| | **symptomatic** | 50 (5.3) | 82 (15.5) |
| | **missing** | 140 (14.9) | 35 (6.6) |
| **Liver metastases, n (%)** | **absent** | 638 (67.7) | 301 (56.9) |
| | **present** | 177 (18.8) | 202 (38.2) |
| | **missing** | 127 (13.5) | 26 (4.9) |
| **LDH, n (%)** | **normal** | 749 (79.5) | 264 (49.9) |
| | **1-2x ULN** | 157 (16.7) | 173 (32.7) |
| | **>2x ULN** | 25 (2.7) | 85 (16.1) |
| | **missing** | 11 (1.2) | 7 (1.3) |
| **Number of affected organs, n (%)** | **<3** | 652 (69.2) | 234 (44.2) |
| | **>2** | 290 (30.8) | 295 (55.8) |
| **Body Mass Index, n (%)** | **underweight** | 15 (1.6) | 6 (1.1) |
| | **normal** | 365 (38.7) | 239 (45.2) |
| | **overweight** | 380 (40.3) | 206 (38.9) |
| | **obese** | 182 (19.3) | 78 (14.7) |
| **Skeletal Muscle Index, median [Q1,Q3]** | | 0.0 [-0.7,0.7] | -0.0 [-0.7,0.5] |
| **Skeletal Muscle Density, median [Q1,Q3]** | | 0.0 [-0.7,0.6] | 0.3 [-0.5,0.8] |
| **Skeletal Muscle Gauge, median [Q1,Q3]** | | -0.0 [-0.7,0.6] | 0.2 [-0.5,0.8] |
| **Subcutaneous Adipose Tissue Index, median [Q1,Q3]** | | -0.2 [-0.6,0.4] | -0.2 [-0.7,0.4] |
| **Visceral Adipose Tissue Index, median [Q1,Q3]** | | -0.1 [-0.7,0.7] | -0.3 [-0.9,0.5] |
| **Median overall survival (months)** | | 34.2 | 37.7 |
| **Median progression-free survival (months)** | | 9.9 | 6.5 |
| **Median melanoma-specific survival (months)** | | 66.5 | 51.2 |

**Supplementary Table 3 - Characteristics and outcomes of patients who underwent contrast-enhanced versus non-contrast CT**

| | | Contrast-enhanced | No contrast |
|---|---|---|---|
| **n** | | 860 | 611 |
| **Age, mean (SD)** | | 64.8 (12.6) | 65.4 (13.5) |
| **Sex, n (%)** | **Female** | 338 (39.3) | 241 (39.4) |
| | **Male** | 522 (60.7) | 370 (60.6) |
| **Therapy, n (%)** | **Anti-PD1** | 538 (62.6) | 404 (66.1) |
| | **Ipilimumab & Nivolumab** | 322 (37.4) | 207 (33.9) |
| **Stage, n (%)** | **IIIC** | 61 (7.1) | 70 (11.5) |
| | **IV M1a** | 67 (7.8) | 63 (10.3) |
| | **IV M1b** | 136 (15.8) | 81 (13.3) |
| | **IV M1c** | 379 (44.1) | 260 (42.6) |
| | **IV M1d** | 211 (24.5) | 133 (21.8) |
| | **missing** | 6 (0.7) | 4 (0.7) |
| **ECOG performance status, n (%)** | **0** | 461 (53.6) | 337 (55.2) |
| | **1** | 284 (33.0) | 205 (33.6) |
| | **2-4** | 75 (8.7) | 35 (5.7) |
| | **missing** | 40 (4.7) | 34 (5.6) |
| **Brain metastases, n (%)** | **absent** | 559 (65.0) | 393 (64.3) |
| | **asymptomatic** | 117 (13.6) | 95 (15.5) |
| | **symptomatic** | 94 (10.9) | 38 (6.2) |
| | **missing** | 90 (10.5) | 85 (13.9) |
| **Liver metastases, n (%)** | **absent** | 547 (63.6) | 392 (64.2) |
| | **present** | 242 (28.1) | 137 (22.4) |
| | **missing** | 71 (8.3) | 82 (13.4) |
| **LDH, n (%)** | **normal** | 576 (67.0) | 437 (71.5) |
| | **1-2x ULN** | 204 (23.7) | 126 (20.6) |
| | **>2x ULN** | 68 (7.9) | 42 (6.9) |
| | **missing** | 12 (1.4) | 6 (1.0) |
| **Number of affected organs, n (%)** | **<3** | 496 (57.7) | 390 (63.8) |
| | **>2** | 364 (42.3) | 221 (36.2) |
| **Body Mass Index, n (%)** | **underweight** | 15 (1.7) | 6 (1.0) |
| | **normal** | 351 (40.8) | 253 (41.4) |
| | **overweight** | 336 (39.1) | 250 (40.9) |
| | **obese** | 158 (18.4) | 102 (16.7) |
| **Skeletal Muscle Index, median [Q1,Q3]** | | -0.0 [-0.7,0.6] | 0.1 [-0.6,0.7] |
| **Skeletal Muscle Density, median [Q1,Q3]** | | 0.1 [-0.7,0.7] | 0.1 [-0.6,0.7] |
| **Skeletal Muscle Gauge, median [Q1,Q3]** | | 0.1 [-0.6,0.7] | 0.0 [-0.6,0.7] |
| **Subcutaneous Adipose Tissue Index, median [Q1,Q3]** | | -0.2 [-0.6,0.4] | -0.2 [-0.7,0.4] |
| **Visceral Adipose Tissue Index, median [Q1,Q3]** | | -0.1 [-0.8,0.7] | -0.2 [-0.8,0.5] |
| **Median overall survival (months)** | | 30.4 | 43.1 |
| **Median progression-free survival (months)** | | 8.3 | 9.8 |
| **Median melanoma-specific survival (months)** | | not reached | 66.5 |

**Supplementary Table 4 - Characteristics and outcomes of patients according to BMI subgroup**

| | | underweight | normal | overweight | obese |
|---|---|---|---|---|---|
| **n** | | 21 | 604 | 586 | 260 |
| **Age, mean (SD)** | | 61.6 (17.2) | 65.2 (14.2) | 65.3 (12.2) | 64.5 (11.4) |
| **Sex, n (%)** | **Female** | 17 (81.0) | 267 (44.2) | 188 (32.1) | 107 (41.2) |
| | **Male** | 4 (19.0) | 337 (55.8) | 398 (67.9) | 153 (58.8) |
| **Therapy, n (%)** | **Anti-PD1** | 15 (71.4) | 365 (60.4) | 380 (64.8) | 182 (70.0) |
| | **Ipilimumab & Nivolumab** | 6 (28.6) | 239 (39.6) | 206 (35.2) | 78 (30.0) |
| **Scan type, n (%)** | **Contrast-enhanced** | 15 (71.4) | 351 (58.1) | 336 (57.3) | 158 (60.8) |
| | **No contrast** | 6 (28.6) | 253 (41.9) | 250 (42.7) | 102 (39.2) |
| **Stage, n (%)** | **IIIC** | 2 (9.5) | 57 (9.4) | 39 (6.7) | 33 (12.7) |
| | **IV M1a** | 1 (4.8) | 37 (6.1) | 63 (10.8) | 29 (11.2) |
| | **IV M1b** | 2 (9.5) | 78 (12.9) | 88 (15.0) | 49 (18.8) |
| | **IV M1c** | 8 (38.1) | 286 (47.4) | 242 (41.3) | 103 (39.6) |
| | **IV M1d** | 7 (33.3) | 146 (24.2) | 148 (25.3) | 43 (16.5) |
| | **missing** | 1 (4.8) | | 6 (1.0) | 3 (1.2) |
| **ECOG performance status, n (%)** | **0** | 11 (52.4) | 321 (53.1) | 334 (57.0) | 132 (50.8) |
| | **1** | 6 (28.6) | 208 (34.4) | 175 (29.9) | 100 (38.5) |
| | **2-4** | 3 (14.3) | 53 (8.8) | 38 (6.5) | 16 (6.2) |
| | **missing** | 1 (4.8) | 22 (3.6) | 39 (6.7) | 12 (4.6) |
| **Brain metastases, n (%)** | **absent** | 12 (57.1) | 388 (64.2) | 375 (64.0) | 177 (68.1) |
| | **asymptomatic** | 6 (28.6) | 84 (13.9) | 96 (16.4) | 26 (10.0) |
| | **symptomatic** | 1 (4.8) | 62 (10.3) | 52 (8.9) | 17 (6.5) |
| | **missing** | 2 (9.5) | 70 (11.6) | 63 (10.8) | 40 (15.4) |
| **Liver metastases, n (%)** | **absent** | 9 (42.9) | 368 (60.9) | 384 (65.5) | 178 (68.5) |
| | **present** | 10 (47.6) | 175 (29.0) | 151 (25.8) | 43 (16.5) |
| | **missing** | 2 (9.5) | 61 (10.1) | 51 (8.7) | 39 (15.0) |
| **LDH, n (%)** | **normal** | 10 (47.6) | 410 (67.9) | 412 (70.3) | 181 (69.6) |
| | **1-2x ULN** | 8 (38.1) | 138 (22.8) | 123 (21.0) | 61 (23.5) |
| | **>2x ULN** | 3 (14.3) | 51 (8.4) | 42 (7.2) | 14 (5.4) |
| | **missing** | | 5 (0.8) | 9 (1.5) | 4 (1.5) |
| **Number of affected organs, n (%)** | **<3** | 12 (57.1) | 349 (57.8) | 355 (60.6) | 170 (65.4) |
| | **>2** | 9 (42.9) | 255 (42.2) | 231 (39.4) | 90 (34.6) |
| **Skeletal Muscle Index, median [Q1,Q3]** | | -1.5 [-1.7,-1.2] | -0.5 [-1.0,0.1] | 0.2 [-0.3,0.7] | 0.8 [0.1,1.5] |
| **Skeletal Muscle Density, median [Q1,Q3]** | | 0.6 [-0.0,1.1] | 0.4 [-0.3,0.9] | 0.1 [-0.7,0.6] | -0.6 [-1.3,0.1] |
| **Skeletal Muscle Gauge, median [Q1,Q3]** | | 0.1 [-0.3,0.5] | 0.2 [-0.4,0.8] | 0.1 [-0.6,0.7] | -0.5 [-1.2,0.3] |
| **Subcutaneous Adipose Tissue Index, median [Q1,Q3]** | | -1.3 [-1.4,-0.9] | -0.6 [-0.9,-0.2] | -0.0 [-0.4,0.4] | 1.0 [0.2,1.8] |
| **Visceral Adipose Tissue Index, median [Q1,Q3]** | | -1.3 [-1.4,-1.1] | -0.7 [-1.1,-0.2] | 0.1 [-0.4,0.7] | 0.9 [0.3,1.6] |
| **Median overall survival (months)** | | 11.6 | 38.2 | 41.4 | 32.9 |
| **Median progression-free survival (months)** | | 3 | 8.7 | 9.8 | 7.7 |
| **Median melanoma-specific survival (months)** | | 18.5 | not reached | not reached | 66.5 |

### Supplementary Table 5 - Pearson's correlation coefficients for body composition metrics

| | BMI | SMI | SMD | SMG | SATI | VATI |
|---|---|---|---|---|---|---|
| **BMI** | r=1.00 (p=0.00) | r=0.54 (p=0.00) | r=-0.34 (p=0.00) | r=-0.23 (p=0.00) | r=0.69 (p=0.00) | r=0.62 (p=0.00) |
| **SMI** | | r=1.00 (p=0.00) | r=-0.14 (p=0.00) | r=0.07 (p=0.01) | r=0.18 (p=0.00) | r=0.61 (p=0.00) |
| **SMD** | | | r=1.00 (p=0.00) | r=0.96 (p=0.00) | r=-0.41 (p=0.00) | r=-0.38 (p=0.00) |
| **SMG** | | | | r=1.00 (p=0.00) | r=-0.38 (p=0.00) | r=-0.25 (p=0.00) |
| **SATI** | | | | | r=1.00 (p=0.00) | r=0.33 (p=0.00) |
| **VATI** | | | | | | r=1.00 (p=0.00) |

Abbreviations: BMI=body mass index, SMI=skeletal muscle index, SMD=skeletal muscle density, SMG=skeletal muscle gauge, SATI=subcutaneous adipose tissue index, VATI=visceral adipose tissue index

**Supplementary Table 6 - Univariate and multivariate Cox proportional hazards models for progression free survival in subgroup of patients treated with anti-PD1 (N=942)**

| | | Univariate | | | Multivariate* | | |
|---|---|---|---|---|---|---|---|
| | | HR | 95% CI | p-value | HR | 95% CI | p-value |
| Body Mass Index (categorical) | underweight | 1.938 | 1.108 - 3.388 | 0.021 | 1.799 | 1.019 - 3.179 | 0.043 |
| | normal | 1.000 | | | 1.000 | | |
| | overweight | 1.036 | 0.870 - 1.232 | 0.693 | 1.043 | 0.875 - 1.243 | 0.639 |
| | obese | 1.078 | 0.872 - 1.334 | 0.487 | 1.174 | 0.946 - 1.457 | 0.147 |
| Body Mass Index (continuous) | | 1.004 | 0.987 - 1.021 | 0.633 | 1.011 | 0.994 - 1.029 | 0.189 |
| Skeletal Muscle Index | | 1.067 | 0.990 - 1.150 | 0.089 | 1.110 | 1.028 - 1.198 | 0.008 |
| Skeletal Muscle Density | | 0.955 | 0.885 - 1.031 | 0.237 | 0.958 | 0.885 - 1.037 | 0.289 |
| Skeletal Muscle Gauge | | 0.985 | 0.913 - 1.063 | 0.700 | 0.995 | 0.919 - 1.078 | 0.911 |
| Subcutaneous Adipose Tissue Index | | 0.969 | 0.894 - 1.050 | 0.444 | 0.982 | 0.904 - 1.065 | 0.655 |
| Visceral Adipose Tissue Index | | 1.085 | 1.008 - 1.168 | 0.030 | 1.092 | 1.012 - 1.178 | 0.024 |

*Corrected for age, sex, serum lactate dehydrogenase, presence of brain metastases (absent vs. asymptomatic vs. symptomatic) and liver metastases, Eastern Cooperative Oncology group performance status and number of affected organs. Abbreviations: HR=Hazard Rate Ratio, CI=Confidence Interval

**Supplementary Table 7 - Univariate and multivariate Cox proportional hazards models for overall survival in subgroup of patients treated with anti-PD1 (N=942)**

| | | Univariate | | | Multivariate* | | |
|---|---|---|---|---|---|---|---|
| | | HR | 95% CI | p-value | HR | 95% CI | p-value |
| Body Mass Index (categorical) | underweight | 1.190 | 0.609 - 2.325 | 0.610 | 0.974 | 0.489 - 1.941 | 0.940 |
| | normal | 1.000 | | | 1.000 | | |
| | overweight | 0.931 | 0.757 - 1.144 | 0.495 | 0.986 | 0.800 - 1.215 | 0.892 |
| | obese | 0.933 | 0.726 - 1.200 | 0.592 | 1.085 | 0.838 - 1.404 | 0.536 |
| Body Mass Index (continuous) | | 0.989 | 0.969 - 1.009 | 0.275 | 1.003 | 0.983 - 1.023 | 0.763 |
| Skeletal Muscle Index | | 1.028 | 0.942 - 1.123 | 0.535 | 1.110 | 1.014 - 1.216 | 0.024 |
| Skeletal Muscle Density | | 0.841 | 0.771 - 0.917 | 0.000 | 0.857 | 0.782 - 0.939 | 0.001 |
| Skeletal Muscle Gauge | | 0.857 | 0.787 - 0.934 | 0.000 | 0.883 | 0.806 - 0.968 | 0.008 |
| Subcutaneous Adipose Tissue Index | | 0.893 | 0.808 - 0.986 | 0.026 | 0.918 | 0.831 - 1.013 | 0.090 |
| Visceral Adipose Tissue Index | | 1.131 | 1.038 - 1.233 | 0.005 | 1.154 | 1.054 - 1.262 | 0.002 |

*Corrected for age, sex, serum lactate dehydrogenase, presence of brain metastases (absent vs. asymptomatic vs. symptomatic) and liver metastases, Eastern Cooperative Oncology group performance status and number of affected organs. Abbreviations: HR=Hazard Rate Ratio, CI=Confidence Interval

**Supplementary Table 8 - Univariate and multivariate Cox proportional hazards models for progression free survival in subgroup of patients treated with ipilimumab plus nivolumab (N=529)**

| | | Univariate | | | Multivariate* | | |
|---|---|---|---|---|---|---|---|
| | | **HR** | **95% CI** | **p-value** | **HR** | **95% CI** | **p-value** |
| **Body Mass Index (categorical)** | **underweight** | 1.954 | 0.724 - 5.275 | 0.187 | 1.563 | 0.565 - 4.323 | 0.390 |
| | **normal** | 1.000 | | | 1.000 | | |
| | **overweight** | 0.906 | 0.715 - 1.150 | 0.418 | 0.875 | 0.687 - 1.114 | 0.281 |
| | **obese** | 0.970 | 0.701 - 1.342 | 0.855 | 0.960 | 0.689 - 1.336 | 0.807 |
| **Body Mass Index (continuous)** | | 0.978 | 0.952 - 1.004 | 0.095 | 0.978 | 0.952 - 1.004 | 0.098 |
| **Skeletal Muscle Index** | | 0.954 | 0.854 - 1.067 | 0.413 | 0.976 | 0.871 - 1.095 | 0.682 |
| **Skeletal Muscle Density** | | 0.997 | 0.891 - 1.115 | 0.957 | 1.046 | 0.934 - 1.173 | 0.434 |
| **Skeletal Muscle Gauge** | | 0.983 | 0.877 - 1.101 | 0.763 | 1.038 | 0.924 - 1.166 | 0.528 |
| **Subcutaneous Adipose Tissue Index** | | 0.944 | 0.853 - 1.044 | 0.260 | 0.965 | 0.872 - 1.067 | 0.484 |
| **Visceral Adipose Tissue Index** | | 1.036 | 0.927 - 1.159 | 0.532 | 1.026 | 0.916 - 1.150 | 0.657 |

*Corrected for age, sex, serum lactate dehydrogenase, presence of brain metastases (absent vs. asymptomatic vs. symptomatic) and liver metastases, Eastern Cooperative Oncology group performance status and number of affected organs. Abbreviations: HR=Hazard Rate Ratio, CI=Confidence Interval

**Supplementary Table 9 - Univariate and multivariate Cox proportional hazards models for overall survival in subgroup of patients treated with ipilimumab plus nivolumab (N=529)**

| | | Univariate | | | Multivariate* | | |
|---|---|---|---|---|---|---|---|
| | | **HR** | **95% CI** | **p-value** | **HR** | **95% CI** | **p-value** |
| **Body Mass Index (categorical)** | **underweight** | 3.194 | 1.175 - 8.688 | 0.024 | 2.203 | 0.791 - 6.136 | 0.132 |
| | **normal** | 1.000 | | | 1.000 | | |
| | **overweight** | 0.971 | 0.727 - 1.296 | 0.840 | 0.988 | 0.737 - 1.324 | 0.933 |
| | **obese** | 1.142 | 0.778 - 1.677 | 0.499 | 1.199 | 0.809 - 1.776 | 0.368 |
| **Body Mass Index (continuous)** | | 1.001 | 0.971 - 1.032 | 0.939 | 1.009 | 0.979 - 1.039 | 0.569 |
| **Skeletal Muscle Index** | | 0.997 | 0.872 - 1.140 | 0.968 | 1.063 | 0.926 - 1.219 | 0.386 |
| **Skeletal Muscle Density** | | 0.888 | 0.778 - 1.014 | 0.080 | 0.905 | 0.791 - 1.034 | 0.144 |
| **Skeletal Muscle Gauge** | | 0.883 | 0.771 - 1.012 | 0.075 | 0.917 | 0.798 - 1.055 | 0.227 |
| **Subcutaneous Adipose Tissue Index** | | 1.000 | 0.889 - 1.123 | 0.994 | 1.043 | 0.930 - 1.169 | 0.474 |
| **Visceral Adipose Tissue Index** | | 1.173 | 1.032 - 1.333 | 0.015 | 1.209 | 1.063 - 1.376 | 0.004 |

*Corrected for age, sex, serum lactate dehydrogenase, presence of brain metastases (absent vs. asymptomatic vs. symptomatic) and liver metastases, Eastern Cooperative Oncology group performance status and number of affected organs. Abbreviations: HR=Hazard Rate Ratio, CI=Confidence Interval

**Supplementary Table 10 - Univariate and multivariate Cox proportional hazards models for progression free survival in subgroup of patients who underwent a contrast-enhanced CT scan (N=860)**

| | | Univariate | | | Multivariate* | | |
|---|---|---|---|---|---|---|---|
| | | HR | 95% CI | p-value | HR | 95% CI | p-value |
| Body Mass Index (categorical) | underweight | 2.061 | 1.177 - 3.607 | 0.012 | 2.214 | 1.257 - 3.897 | 0.006 |
| | normal | 1.000 | | | 1.000 | | |
| | overweight | 1.007 | 0.839 - 1.207 | 0.944 | 1.009 | 0.840 - 1.212 | 0.925 |
| | obese | 0.927 | 0.736 - 1.169 | 0.523 | 1.015 | 0.803 - 1.283 | 0.900 |
| Body Mass Index (continuous) | | 0.986 | 0.968 - 1.005 | 0.140 | 0.992 | 0.974 - 1.011 | 0.412 |
| Skeletal Muscle Index | | 0.978 | 0.902 - 1.061 | 0.595 | 1.017 | 0.936 - 1.105 | 0.685 |
| Skeletal Muscle Density | | 0.958 | 0.884 - 1.039 | 0.300 | 0.957 | 0.881 - 1.040 | 0.303 |
| Skeletal Muscle Gauge | | 0.975 | 0.899 - 1.057 | 0.541 | 0.979 | 0.901 - 1.064 | 0.621 |
| Subcutaneous Adipose Tissue Index | | 0.972 | 0.897 - 1.054 | 0.496 | 0.988 | 0.911 - 1.071 | 0.766 |
| Visceral Adipose Tissue Index | | 1.007 | 0.932 - 1.088 | 0.863 | 1.029 | 0.950 - 1.113 | 0.487 |

*Corrected for age, sex, serum lactate dehydrogenase, presence of brain metastases (absent vs. asymptomatic vs. symptomatic) and liver metastases, Eastern Cooperative Oncology group performance status and number of affected organs. Abbreviations: HR=Hazard Rate Ratio, CI=Confidence Interval

**Supplementary Table 11 - Univariate and multivariate Cox proportional hazards models for overall survival in subgroup of patients who underwent a contrast-enhanced CT scan (N=860)**

| | | Univariate | | | Multivariate* | | |
|---|---|---|---|---|---|---|---|
| | | HR | 95% CI | p-value | HR | 95% CI | p-value |
| Body Mass Index (categorical) | underweight | 1.582 | 0.809 - 3.093 | 0.181 | 1.914 | 0.974 - 3.759 | 0.060 |
| | normal | 1.000 | | | 1.000 | | |
| | overweight | 1.001 | 0.807 - 1.241 | 0.994 | 1.066 | 0.858 - 1.326 | 0.562 |
| | obese | 0.877 | 0.665 - 1.156 | 0.352 | 1.056 | 0.795 - 1.402 | 0.708 |
| Body Mass Index (continuous) | | 0.989 | 0.968 - 1.011 | 0.327 | 1.003 | 0.981 - 1.026 | 0.772 |
| Skeletal Muscle Index | | 1.002 | 0.911 - 1.103 | 0.965 | 1.069 | 0.969 - 1.180 | 0.184 |
| Skeletal Muscle Density | | 0.863 | 0.785 - 0.949 | 0.003 | 0.847 | 0.768 - 0.934 | 0.001 |
| Skeletal Muscle Gauge | | 0.878 | 0.799 - 0.965 | 0.007 | 0.872 | 0.790 - 0.962 | 0.007 |
| Subcutaneous Adipose Tissue Index | | 0.945 | 0.856 - 1.043 | 0.264 | 0.980 | 0.889 - 1.081 | 0.692 |
| Visceral Adipose Tissue Index | | 1.086 | 0.993 - 1.188 | 0.070 | 1.138 | 1.038 - 1.247 | 0.006 |

*Corrected for age, sex, serum lactate dehydrogenase, presence of brain metastases (absent vs. asymptomatic vs. symptomatic) and liver metastases, Eastern Cooperative Oncology group performance status and number of affected organs. Abbreviations: HR=Hazard Rate Ratio, CI=Confidence Interval

**Supplementary Table 12 - Univariate and multivariate Cox proportional hazards models for progression free survival in subgroup of patients who underwent a non-contrast CT scan (N=611)**

| | | Univariate | | | Multivariate* | | |
|---|---|---|---|---|---|---|---|
| | | HR | 95% CI | p-value | HR | 95% CI | p-value |
| Body Mass Index (categorical) | underweight | 1.342 | 0.497 - 3.619 | 0.562 | 1.052 | 0.378 - 2.925 | 0.922 |
| | normal | 1.000 | | | 1.000 | | |
| | overweight | 0.958 | 0.769 - 1.193 | 0.700 | 0.973 | 0.778 - 1.216 | 0.809 |
| | obese | 1.220 | 0.927 - 1.606 | 0.156 | 1.207 | 0.912 - 1.596 | 0.189 |
| Body Mass Index (continuous) | | 1.009 | 0.987 - 1.031 | 0.421 | 1.010 | 0.989 - 1.032 | 0.345 |
| Skeletal Muscle Index | | 1.111 | 1.005 - 1.228 | 0.041 | 1.137 | 1.028 - 1.256 | 0.012 |
| Skeletal Muscle Density | | 0.986 | 0.893 - 1.089 | 0.777 | 1.026 | 0.924 - 1.140 | 0.627 |
| Skeletal Muscle Gauge | | 0.999 | 0.904 - 1.105 | 0.989 | 1.049 | 0.944 - 1.165 | 0.379 |
| Subcutaneous Adipose Tissue Index | | 0.943 | 0.854 - 1.041 | 0.245 | 0.958 | 0.870 - 1.056 | 0.387 |
| Visceral Adipose Tissue Index | | 1.172 | 1.058 - 1.298 | 0.003 | 1.164 | 1.049 - 1.293 | 0.005 |

*Corrected for age, sex, serum lactate dehydrogenase, presence of brain metastases (absent vs. asymptomatic vs. symptomatic) and liver metastases, Eastern Cooperative Oncology group performance status and number of affected organs. Abbreviations: HR=Hazard Rate Ratio, CI=Confidence Interval

**Supplementary Table 13 - Univariate and multivariate Cox proportional hazards models for overall survival in subgroup of patients who underwent a non-contrast CT scan (N=611)**

| | | Univariate | | | Multivariate* | | |
|---|---|---|---|---|---|---|---|
| | | HR | 95% CI | p-value | HR | 95% CI | p-value |
| Body Mass Index (categorical) | underweight | 1.315 | 0.485 - 3.567 | 0.591 | 0.721 | 0.255 - 2.043 | 0.539 |
| | normal | 1.000 | | | 1.000 | | |
| | overweight | 0.879 | 0.673 - 1.148 | 0.345 | 0.915 | 0.698 - 1.199 | 0.519 |
| | obese | 1.196 | 0.868 - 1.650 | 0.275 | 1.244 | 0.894 - 1.729 | 0.196 |
| Body Mass Index (continuous) | | 0.998 | 0.972 - 1.024 | 0.881 | 1.006 | 0.982 - 1.031 | 0.616 |
| Skeletal Muscle Index | | 1.044 | 0.930 - 1.172 | 0.462 | 1.116 | 0.995 - 1.252 | 0.062 |
| Skeletal Muscle Density | | 0.857 | 0.767 - 0.958 | 0.007 | 0.895 | 0.795 - 1.008 | 0.068 |
| Skeletal Muscle Gauge | | 0.860 | 0.770 - 0.961 | 0.008 | 0.909 | 0.806 - 1.024 | 0.117 |
| Subcutaneous Adipose Tissue Index | | 0.922 | 0.817 - 1.041 | 0.190 | 0.967 | 0.864 - 1.082 | 0.560 |
| Visceral Adipose Tissue Index | | 1.220 | 1.084 - 1.373 | 0.001 | 1.249 | 1.106 - 1.410 | 0.000 |

*Corrected for age, sex, serum lactate dehydrogenase, presence of brain metastases (absent vs. asymptomatic vs. symptomatic) and liver metastases, Eastern Cooperative Oncology group performance status and number of affected organs. Abbreviations: HR=Hazard Rate Ratio, CI=Confidence Interval
